# Large-scale Testing for SARS-CoV-2 using Tapestry Pooling

**DOI:** 10.1101/2020.10.09.20209742

**Authors:** Anirudh Chakravarthy, Srikar Krishna, Sumana Ghosh, Ajay Tomar, Sriram Varahan, Ajit Rajwade, Sabyasachi Ghosh, Nimay Gupta, Rishi Agarwal, Himanshu Payal, Prantik Chakraborty, Krishna Vishal Vemula, Akanksha Vyas, Ritesh Goru, Sandeep Krishna, Dasaradhi Palakodeti, Manoj Gopalkrishnan

## Abstract

We have previously described Tapestry Pooling, a scheme to enhance the capacity of RT-qPCR testing, and provided experimental evidence with spiked synthetic RNA to show that it can help to scale testing and restart the economy. Here we report on validation studies with Covid19 patient samples for the Tapestry Pooling scheme with prevalence in the range of 1% to 2%. We pooled RNA extracted from patient samples that were previously tested for Covid19, sending each sample to three pools. Following three different pooling schemes, we pipetted 320 samples into 48 pools with pool size of 20 at prevalence rate of 1.6%, 500 samples into 60 pools with pool size of 25 at prevalence rate of 2%, and 961 samples into 93 pools with pool size of 31 at prevalence rate of 1%. Of the 191 RT-qPCR experiments that we performed, only one pool was incorrect (false negative). Our recovery algorithm correctly called results for the individual samples, with a 100% sensitivity and a 99.9% specificity, with only one false positive across all the 1,781 blinded results required to be called. We show up to 10X savings in number of tests required at a range of prevalence rates and pool sizes. These experiments establish that Tapestry Pooling is robust enough to handle the diversity of sample constitutions and viral loads seen in real-world samples.

## Introduction

The SARS-CoV-2 virus is now active in all populated parts of the world, with the cumulative number of infections globally being estimated over 33 million as of September 2020. While the virus was initially met with large-scale lockdowns, these have largely been lifted and replaced by smaller-scale lockdowns around known clusters of infection. Several establishments such as schools, offices, airports, hospitals, shopping malls remain restricted or working at less than full capacity. Communities are forced to trade off economic distress against morbidity and mortality. Such decisions would be eased with the availability of an accurate and affordable method for large-scale and relatively rapid testing of individuals for SARS-CoV-2.

Pooling samples enhances the capacity of qRT-PCR assays to meet the growing demand for testing. If a pool tests negative, all samples in that pool are declared negative. When the prevalence of infection is low, many pools turn out to be negative. Each test gives results for multiple samples, allowing available resources to be deployed to perform more tests. One such two-round pooling strategy that has been used in India as well as in Israel, Germany, China, and the USA has been to take pools of 5 samples each. If a pool tests negative by qRT-PCR, all samples participating in that pool are declared negative. For each positive pool, every sample within that pool is individually re-tested to determine if it is positive or negative for the virus. We term this “simple pooling”.

We have previously proposed an algorithmic method of pooling, known as Tapestry Pooling, to achieve substantial reductions of the number of tests required per sample, compared to simple pooling (1, 2). Tapestry Pooling identifies positives in a single round of testing. It can give savings at prevalence as high as 20%. At very low prevalence, it can deliver more than 10 results per test. These advantages make Tapestry Pooling an attractive alternative to simple pooling. In Tapestry Pooling, each sample goes to three pools while ensuring that any two samples share at most one pool. Our novel reconstruction algorithm recovers the presence or absence as well as amount of the virus in each individual sample from a *single round* of qRT-PCR results of the pools. We have combined ideas from the fields of Combinatorial Group Testing, Compressed Sensing and Sparse Regression to devise the algorithm (1).

We previously validated Tapestry Pooling in experiments where fake positive samples were created by spiking with synthetic template RNA (2). We have also argued in Ghosh *et al* that Tapestry Pooling has various advantages over other algorithmic pooling schemes (3). In this paper, we describe experiments on Covid19 patient samples and show that Tapestry Pooling works well for population sizes ranging from 300 to 1000 when prevalence rates are in the range of 1-2% and pool sizes range from 20 to 31. Elsewhere we will report the validation of Tapestry Pooling for scenarios where the prevalence rate is as high as 15-20% (manuscript under preparation).

## Methods

### Study Design

We performed three case studies. Each case study was implemented as follows:

1. We chose the number of individuals and the prevalence rate to be tested.
2. Given these numbers and without knowing precisely which samples will be positive or negative, we chose a **pooling scheme** that determines which samples participate in which pools. The pooling schemes can be found in Supplementary Table S1.
3. The experimental team chose which samples were negative and which were positive, in accordance with the prevalence rate chosen above. Positive samples were chosen randomly from among previously-tested positive patient samples (Supplementary Table S1), so that the variation in their viral loads was indicative of natural variation in viral loads for patient samples arriving at a testing lab.
4. The experimental team then pooled pre-extracted RNA from the samples according to the pooling matrix provided, and conducted qRT-PCR tests on the pools. In each pooling scheme, every sample participated in exactly 3 pools, while the pool sizes ranged from 20 to 31.
5. The C_t_ values obtained from the qRT-PCR runs were reported back to an analysis team. The C_t_ values for the three pooling schemes tested are shown in supplementary table S2a, S2b and S2c. Which individual samples were positive or negative was blinded from the analysis team.
6. The analysis team applied the reconstruction algorithm to the qRT-PCR results. The algorithm classified each individual sample into one of three categories: positive, negative and undetermined.
7. This prediction was tallied with the ground truth, namely which samples were actually positive or negative, to compute the sensitivity and specificity of the method.

The following cases were examined:

1. 320 samples, 48 pools, 5 positives (1.6% prevalence), pool size 20.
2. 500 samples, 60 pools, 10 positives (2% prevalence), pool size 25.
3. 961 samples, 93 pools, 10 positives (1% prevalence), pool size 31.

The pooling schemes were chosen in order to explore population sizes ranging from several hundred to around a thousand keeping prevalence rates in the 1-2% range while allowing all pools and controls to fit within a 96-well plate for a single round of qRT-PCR.

### Sample handling and RNA isolation

All pooling experiments done for this study were performed using RNA isolated from patient nasopharyngeal swab samples. The handling of these samples was done as prescribed by the Indian Council of Medical Research. RNA isolation was done using the Qiagen QIAamp Viral RNA Mini kit. The isolated RNA was tested independently by qRT-PCR in addition to being used for this experimental pooling study.

### Pooling strategy

RNA samples that had been isolated previously from clinical swab samples and stored at −80°C were used for the pooling experiments.

#### 48 pools for 320 samples

315 negative samples and 5 positive samples were chosen for pooling. Each pool contained 20 = 320* 3/48 RNA samples. Since this was the first study we were doing with a large pool size, we were concerned about possible over-dilution of positive RNA samples. So we made provision to reduce dilution by pooling 1µL of each negative sample and 2.5µL of each positive sample. Each pool contained between 20 to 23 µL of pooled RNA that was used for subsequent qRT-PCR testing.

#### 60 pools for 500 samples

490 negative samples and 10 positive samples were selected. Each pool contained 25 = 500*3/60 individual samples. From our previous experiment, we discovered that the dilution of RNA samples due to pooling did not drastically increase the Ct values of positive pools in our qRT-PCR runs. Therefore, we added 1µL of each RNA sample (positive and negative samples) into each of its designated pools. Each pool contained 25µL of pooled RNA that was used for subsequent qRT-PCR testing.

#### 93 pools for 961 samples

Each pool contained a total of 31 = 961*3/93 samples. We chose 10 positive samples and 547 negatives. We identified the pools into which the 10 positive samples were added. These amounted to a total of 23 pools. We prepared these pools by pooling the participating positive samples along with several other negative samples per pool. Pooling was done by adding 1uL of each RNA sample (positive and negative samples) resulting in a total of 31µL of RNA per pool that was used for subsequent qRT-PCR testing. The remaining 70 reactions which contained only negative samples and no positives were simulated using 70 unique negative samples as the template.

### Testing using qRT-PCR

Once the pooling of RNA samples was completed, aliquots from the pools were subjected to qRT-PCR testing for Covid-19. The pools were tested using the MyLab qRT-PCR kit (PathoDetect Covid-19). This kit is a multiplex kit capable of detecting the SARS-Cov2 RdRp and E genes. In order to reduce the dilution factor of the positive RNA samples, we used double the volume of template RNA and half the volume of nuclease free water prescribed by the MyLab kit protocol. All other components of the reaction mixture (primers, probes and RT-PCR master mix) were added exactly as prescribed by the MyLab kit instructions with appropriate control reactions. The reaction compositions for each pooling scheme are shown in the tables below.

The thermocycler was set up as recommended in the kit instructions. Results from the qRT-PCR machine were recorded and submitted to the Tapestry recovery algorithm for identification of Covid19 positive samples.

### Performance Statistics

The Tapestry Pooling reconstruction algorithm classifies samples into three categories: positive, negative, and undetermined. The provision for undetermined samples ensures that even when the number of positives in a batch exceeds the capacity of the pooling scheme, we do not compromise on sensitivity or specificity. Therefore the algorithm’s performance must be judged using sensitivity and specificity, as well as the number of undetermined samples it returns. We calculated **sensitivity** = TP/(TP+FN) and **specificity**=TN/(TN+FP), where TP=number of predicted positives that were true positives, FP=number of predicted positives that were actually negative, TN=number of predicted negatives that were true negatives, and FN=number of predicted negatives that were actually positive.

## Results

Tapestry Pooling was found to work in all three studies with sensitivity of 100% and specificity of 99.9% and only 13 undetermined from 1781 samples tested as shown in Table 2. All pools with positive samples were positive except for one pool in the third study. Note that the results of the algorithm were robust to this false negative. The raw data for the pooling schemes used, the measured Ct values (Supplementary table S2a, S2b and S2c), and the ground truth and predicted positive samples is included in the supplementary information (Supplementary table S3a, S3b and S3c).

**Table 1:**
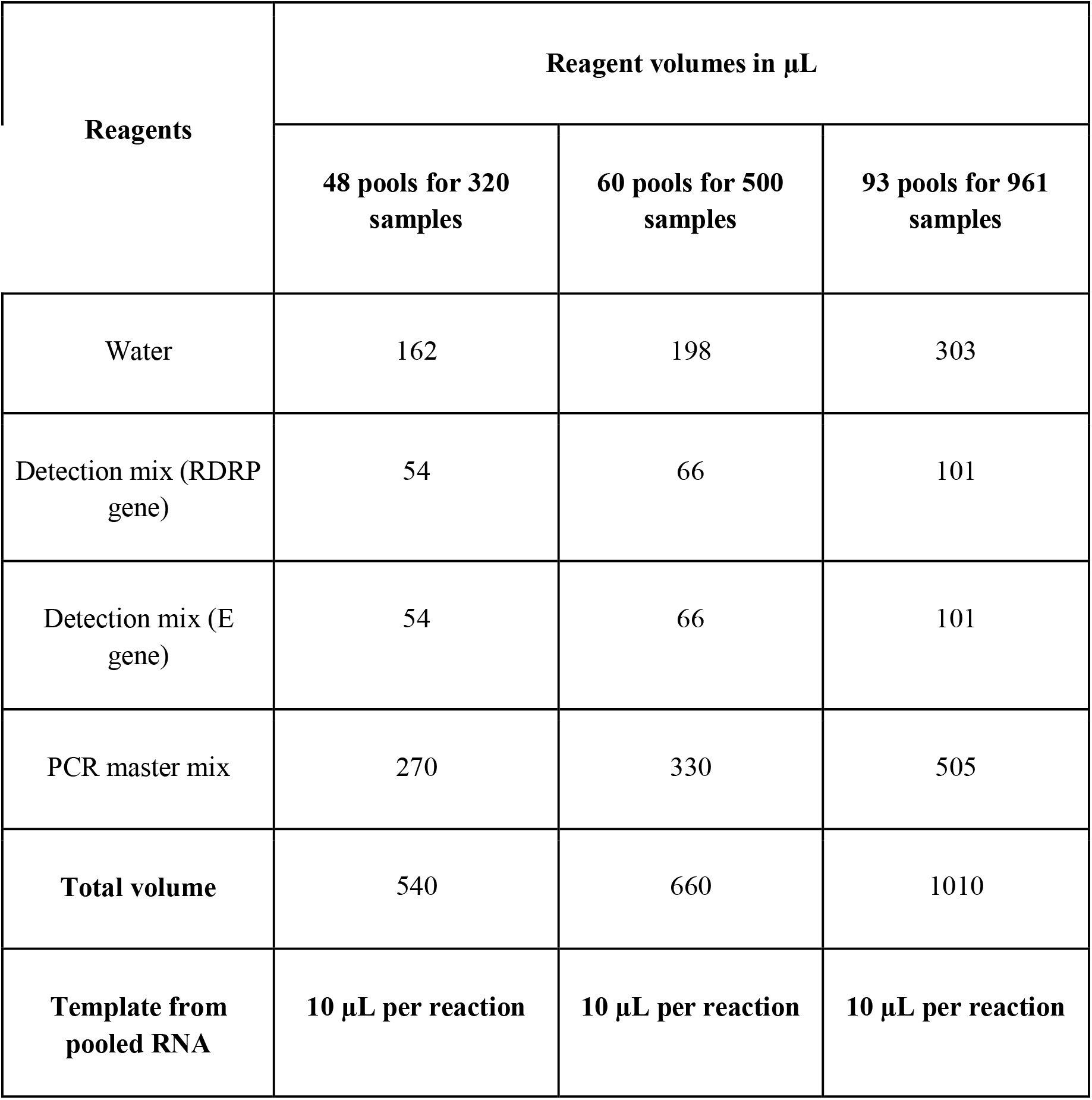
Reaction composition for the pooling schemes used. Total volume per reaction: 20 µL

**Table 2:** High Specificity and Sensitivity of Tapestry Pooling. Tapestry pooling requires substantially fewer tests than simple pooling while preserving high sensitivity and specificity. #Tests represents the number of first round tests plus the number of second round tests. For Tapestry Pooling, this is the number of undetermined samples.

| #Samples | #Positives | Sensitivity | Specificity | #Tests for Tapestry Pooling | #Tests in Simple Pooling |
| --- | --- | --- | --- | --- | --- |
| 320 | 5 | 100% | 100% | 48 + 0 | 64 + 25 |
| 500 | 10 | 100% | 99.8% (1 false positive) | 60 + 10 | 100 + 50 |
| 961 | 10 | 100% | 100% | 93 + 3 | 192 + 50 |

## Discussion

The Tapestry Pooling method is made available as a web app at www.tapestry-pooling.com, making it easy to deploy. The user is assisted to pick a particular matrix size based on their requirements of number of samples to be tested, testing capacity available, prevalence rate expected, and any constraints of pool size for their assay. After pooling and completing the assay, the user enters the Ct value of each pool obtained from the qRT-PCR into the web app. The recovery algorithm solves and returns a result for each sample.

We have demonstrated that Tapestry Pooling works with high sensitivity and specificity as a method for testing large numbers of individuals at low prevalence for SARS-CoV-2. It allows a wide range of pooling schemes to be deployed so that it can be customized over a large range of prevalence rates, number of samples, pool sizes, and number of tests. We have shown that pools of size upto 31 can be managed without false negatives by adjusting the reaction composition, specifically by adding a larger volume of template and less water. This means for example that a positive sample in a pool of size 31 is diluted only 15.5-fold so that the Ct shift is kept within detectable limits. In summary, Tapestry Pooling is a flexible pooling scheme that returns large savings while still maintaining high accuracy.

## Data Availability

All data, appropriately anonymized, is available as a supplementary file to this submission. Our software implementation of the Tapestry Pooling algorithm is being made available as a web app at http://app.tapestry-pooling.com.

https://www.dropbox.com/s/tdjcwcnszkmzps6/Supplementary%20Table%20S1.xlsx?dl=0

## Acknowledgements

Sandeep Krishna thanks the Simons Foundation for funding as well as the Department of Atomic Energy, Government of India under project no. 12-R\&D-TFR-5.04-0800. AR, MG thank SERB MATRICS grants MTR/2019/0006, EMR/2017/004089, MTR/2018/000817, IITB WRCB grant 10013976, and DST-Rakshak grant 10013980. DP acknowledges support by CSR funding from Azim Premji philanthropy, Standard Chartered Bank, and Punjab National Bank,

## Supplementary Data

**Supplementary Table S1: Pooling schemes used**

The 3 pooling schemes tested are provided as Supplementary Excel sheets. In each sheet, the columns represent the pools to be created and list the sample IDs that are grouped together. The positive samples selected for each pooling scheme are marked in red in their respective pools.

**Supplementary Table S2: qRT-PCR results**

The results of the qRT-PCR runs are shown in Table S2 for each pooled test for each of the 3 pooling schemes we studied. The tests for which a cycle threshold (Ct) value was detectable are as shown in the table. The tests not included in the table were undetected in the qRT-PCR reaction. The table shows results for both the RdRp and E genes. The former Ct values were what were fed into the reconstruction algorithm to obtain the results shown in the main text.

**Table S2a:** Cycle threshold values obtained for detectable tests amongst the 48 pooled tests performed with 320 samples.

| Target gene | RdRp gene |  |  |  |  |  |  |  |  |  |  |  |  |  |  |
| --- | --- | --- | --- | --- | --- | --- | --- | --- | --- | --- | --- | --- | --- | --- | --- |
| Test | B1 | B3 | C3 | C11 | D7 | E5 | E7 | F5 | G3 | G5 | G11 | H1 | H3 | H7 | NTC |
| Ct values | 32.84 | 23.69 | 30.35 | 26.72 | 23.55 | 29.25 | 23.43 | 27.08 | 33.21 | 32.88 | 30.35 | 27.15 | 30.23 | 30.18 | NA |

| Target gene | E gene |  |  |  |  |  |  |  |  |  |  |  |  |  |  |
| --- | --- | --- | --- | --- | --- | --- | --- | --- | --- | --- | --- | --- | --- | --- | --- |
| Test | B1 | B3 | C3 | C11 | D7 | E5 | E7 | F5 | G3 | G5 | G11 | H1 | H3 | H7 | NTC |
| Ct values | 32.45 | 22.73 | 29.64 | 26.50 | 22.86 | 28.65 | 22.65 | 26.04 | 32.31 | 31.92 | 29.25 | 26.15 | 29.27 | 29.53 | NA |

**Table S2b:** Cycle threshold values obtained for detectable tests amongst the 60 pooled test performed with 500 samples.

| Target gene | RdRp gene |  |  |  |  |  |  |  |  |  |  |  |  |  |
| --- | --- | --- | --- | --- | --- | --- | --- | --- | --- | --- | --- | --- | --- | --- |
| Test | A1 | A2 | A3 | A6 | A7 | A8 | A12 | B1 | B7 | B9 | B10 | C1 | C2 | C4 |
| Ct values | 31.52 | 29.31 | 31.25 | 28.95 | 25.94 | 23.25 | 25.69 | 41.12 | 25.59 | 28.75 | 35.22 | 37.14 | 34.02 | 23.04 |
| Test | C5 | C8 | C11 | D1 | D3 | D4 | D5 | D6 | D8 | D10 | E3 | E9 | E10 | NTC |
| Ct values | 33.91 | 33.10 | 34.35 | 22.96 | 25.28 | 33.28 | 25.38 | 33.31 | 33.53 | 25.55 | 30.59 | 34.32 | 34.98 | NA |

| Target gene | E gene |  |  |  |  |  |  |  |  |  |  |  |  |  |
| --- | --- | --- | --- | --- | --- | --- | --- | --- | --- | --- | --- | --- | --- | --- |
| Test | A1 | A2 | A3 | A6 | A7 | A8 | A12 | B1 | B7 | B9 | B10 | C1 | C2 | C4 |
| Ct values | 30.83 | 28.21 | 30.34 | 27.98 | 24.50 | 22.09 | 24.79 | 32.78 | 24.26 | 27.56 | 31.66 | 31.07 | 31.40 | 21.69 |
| Test | C5 | C8 | C11 | D1 | D3 | D4 | D5 | D6 | D8 | D10 | E3 | E9 | E10 | NTC |
| Ct values | 31.91 | 31.34 | 30.53 | 21.87 | 24.38 | 31.10 | 24.52 | 31.40 | 31.98 | 24.09 | 30.02 | 31.12 | 31.31 | NA |

**Table S2c:** Cycle threshold values obtained for detectable tests amongst the 93 tests performed with 961 samples.

| Target gene | RdRp gene |  |  |  |  |  |  |  |  |  |  |  |
| --- | --- | --- | --- | --- | --- | --- | --- | --- | --- | --- | --- | --- |
| Test | A12 | B5 | B8 | C1 | C2 | C6 | C8 | C10 | D9 | E1 | E4 | E7 |
| Ct values | 22.34 | 31.92 | 32.59 | 24.76 | 30.89 | 24.63 | 39.53 | 25.44 | 33.89 | 22.46 | 29.47 | 42.58 |
| Test | E10 | E11 | F10 | F11 | G1 | G4 | G8 | G9 | H2 | H4 | NTC |  |
| Ct values | 28.69 | 31.81 | 33.10 | 29.11 | 24.73 | 27.87 | 29.11 | 25.21 | 22.33 | 25.05 | NA |  |

| Target gene | E gene |  |  |  |  |  |  |  |  |  |  |  |
| --- | --- | --- | --- | --- | --- | --- | --- | --- | --- | --- | --- | --- |
| Test | A12 | B5 | B8 | C1 | C2 | C6 | C8 | C10 | D9 | E1 | E4 | E7 |
| Ct values | 23.90 | 33.75 | 33.96 | 26.43 | 33.34 | 26.59 | 35.90 | 26.74 | 33.46 | 23.63 | 32.27 | 35.60 |
| Test | E10 | E11 | F10 | F11 | G1 | G4 | G8 | G9 | H2 | H4 | NTC |  |
| Ct values | 30.29 | 34.94 | 36.86 | 30.38 | 26.66 | 31.06 | 32.62 | 26.49 | 24.27 | 26.28 | NA |  |

**Supplementary Table S3: Prediction results for the pooling scheme**

**Table S3a:** Prediction results for the pooling scheme consisting of 48 pools for 320 samples.

|  |  |
| --- | --- |
| Ground truth (sample IDs that were positive - 5 total) | 46,116,167,238,314 |
| Positives predicted from Tapestry algorithm | 46,116,167,238,314 |
| Number of undetermined from Tapestry algorithm | 0 |
| Number of false positives | 0 |
| Number of false negatives | 0 |

**Table S3b:** Prediction results for the pooling scheme consisting of 60 pools for 500 samples.

|  |  |
| --- | --- |
| Ground truth (sample IDs that were positive - 10 total) | 29, 79, 141, 142, 214, 238, 333, 384, 404, 498 |
| Positives predicted from Tapestry algorithm | 29, 79, 141, 142, 214, 232, 238, 333, 384, 404, 498, |
| Number of undetermined from Tapestry algorithm | 10 |
| Number of false positives | 1 (sample ID 232) |
| Number of false negatives | 0 |

**Table S3c:** Prediction results for the pooling scheme consisting of 93 pools for 961 samples

|  |  |
| --- | --- |
| Ground truth (sample IDs that were positive - 10 total) | 58, 103, 274, 397, 411, 544, 632, 700, 788, 933 |
| Positives predicted from Tapestry algorithm | 58, 103, 274, 397, 411, 544, 632, 700, 788, 933 |
| Number of undetermined from Tapestry algorithm | 3 |
| Number of false positives | 0 |
| Number of false negatives | 0 |

## Notes

### Competing Interest Statement

The authors have declared no competing interest.

### Author Declarations

inStem/IEC-18/01 from Institute for Stem Cell and Regenerative Medicine, Institute Human Ethics Committee dated May 18th 2020.

